# On the effectiveness of COVID-19 restrictions and lockdowns: Pan metron ariston

**DOI:** 10.1101/2021.07.06.21260077

**Authors:** Leonidas Spiliopoulos

## Abstract

I examine the dynamics of confirmed case (and death) growth rates conditional on different levels of severity in implemented NPIs, the mobility of citizens and other non restrictive policies. To account for the endogeneity of many of these variables, and the possibility of correlated latent (unobservable) country characteristics, I estimate a four structural model of the evolution of case growth rates, death growth rates, average changes in mobility and the determination of the severity of NPIs. There are strongly decreasing returns to the stringency of NPIs, especially for extreme lockdowns, as no significant improvement in the main outcome measures is found beyond NPIs corresponding to a Stringency Index range of 51–60 for cases and 41–50 for deaths. A non-restrictive policy of extensive and open testing has half of the impact on pandemic dynamics as the optimal NPIs, with none of the associated social and economic costs resulting from the latter. Decreases in mobility were found to increase, rather than decrease case growth rates, consistent with arguments that within-household transmission–resulting from spending more time at residences due to mobility restrictions–may outweigh the benefits of reduced community transmission. Vaccinations led to a fall in case and death growth rates, however the effect size must be re-evaluated when more data becomes available. Governments conditioned policy choice on recent pandemic dynamics, and were found to de-escalate the associated stringency of implemented NPIs more cautiously than in their escalation, i.e., policy mixes exhibited significant hysteresis. Finally, at least 90% of the maximum effectiveness of NPIs can be achieved by policies with an average Stringency index of 31–40, without restricting internal movement or imposing stay at home measures, and only recommending (not enforcing) closures on workplaces and schools, accompanied by public informational campaigns. Consequently, the positive effects on case and death growth rates of voluntary behavioral changes in response to beliefs about the severity of the pandemic, generally trumped those arising from mandatory behavioral restrictions. The exception being more stringent mandatory restrictions on gatherings and international movement, which were found to be effective. The findings suggest that further work should be directed at re-evaluating the effectiveness of NPIs, particularly towards empirically determining the optimal policy mix and associated stringency of individual NPIs.

## Introduction

The ancient Greek dictum “pan metron ariston” asserts that moderation is best, implying that extreme measures, regardless of which end of the spectrum they are located, are unlikely to be the best policy. With regards to the SARS–CoV-2 pandemic, the one extreme, of no intervention and restrictions was ruled out by most governments early on. The other extreme, of severe restrictions and even complete lockdowns, has not proven to be a bête noire for many governments. Severe lockdowns were imposed during the first wave of the pandemic, when uncertainty regarding the transmission and mortality of Covid-19 was maximal. Such an approach can be justified as a maxmin reaction in the face of uncertain events, minimizing the worst possible outcome. As we learn more about Covid-19 and uncertainty is reduced, better estimates of both the probability distribution and magnitude of possible outcomes can be inferred, allowing for a more nuanced approach aligned with an expected value calculation of costs and benefits. However, severe NPIs remain in place in many countries even during the second and third waves, despite the fact that we are now arguably more informed. This can be seen in the evolution of the stringency of government-imposed NPIs (the Stringency Index, SI, on a scale from 0 to 100) plotted in Figure 1, which includes the median SI value across countries and the 10th and 90th percentiles. Median SI peaked during the first wave in the month of April, reaching a maximum of 84. The median SI then trended slowly downwards and leveled off to a range of 55–60, whilst slowly trending up again as subsequent waves led to a resurgence of the pandemic. As of April 2021, while NPIs had not reached the peak levels of the first wave, the median SI across countries at the last datapoint (14/4/2021) was still relatively high, 64 (10th perc.=31, 90th=81). Another important observation is that after the peak in April 2020, governments have followed increasingly heterogeneous NPIs as the difference in the 10th and 90th percentiles increased from roughly 30 SI points to oscillating around 50 points since late 2020.

**Figure 1:**
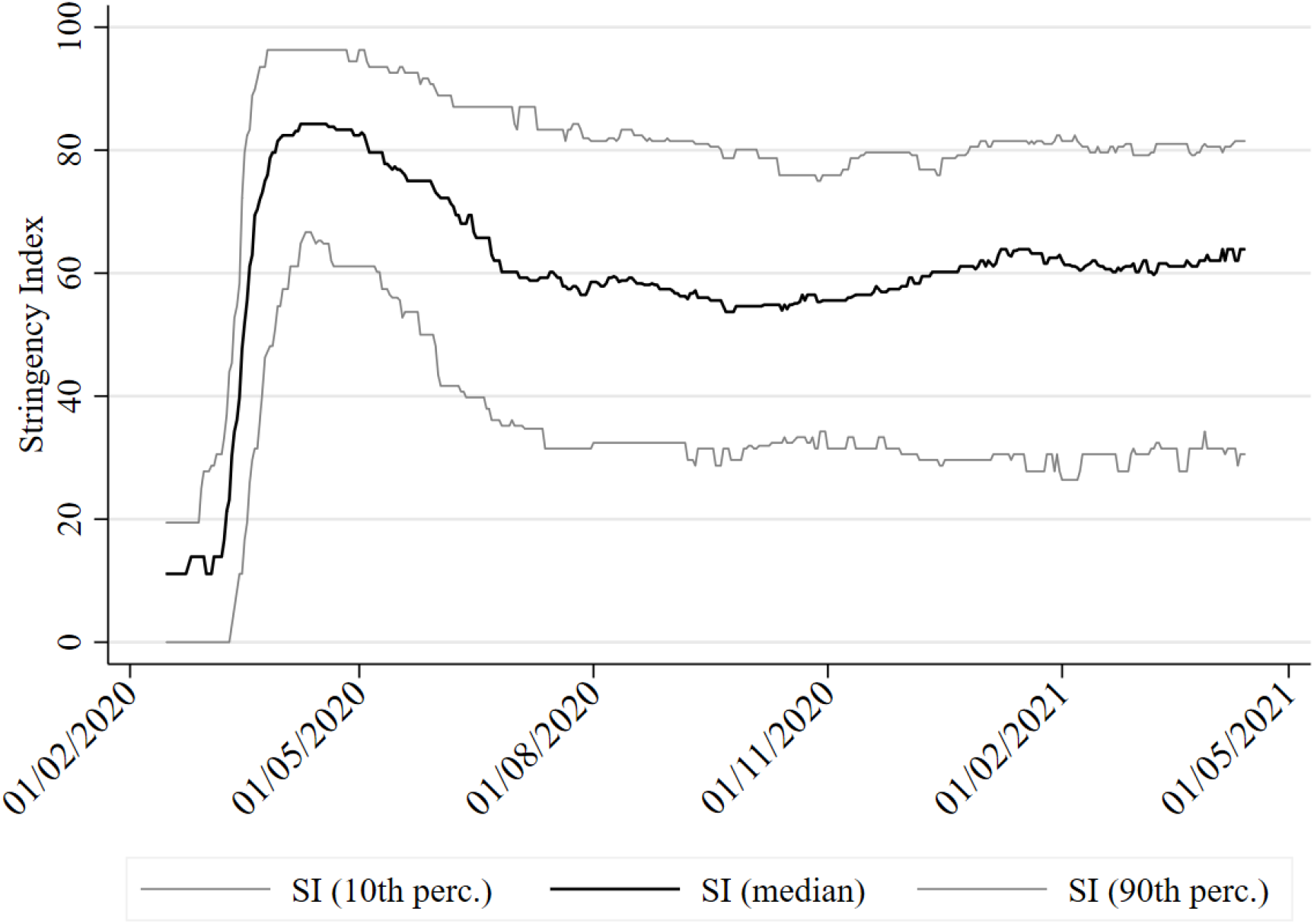
The time evolution of the Stringency Index across countries

The aim of this study is to exploit this divergence in government responses for improved identification of the effects of the strictness of implemented NPIs on the pandemic dynamics. I will extend earlier work based on limited data from the first wave (Haug et al. 2020; Islam et al. 2020; Brauner et al. 2021; Bo et al. 2021) and also deal with certain econometric issues with prior analyses. Does the accumulated data from over a year support the continued imposition of stringent NPIs? I will test the whole spectrum of Covid policies in terms of varying degrees of severity to determine whether metron is indeed ariston. If so, it is crucial that we know where exactly the metron lies within the spectrum of policy responses because restrictions and lockdowns have an immense effect on economic and psychological well being, translating into negative health outcomes in the future (Miles, Stedman and Heald 2020; Susskind and Vines 2020; Altman 2020). The latter are of course difficult to quantify, leading to a propensity to focus instead solely on the positive, immediate effects of NPIs. However, tradeoffs are everywhere and we ignore them at our peril–open debates allowing the free exchange of ideas are of paramount importance (e.g., Melnick and Ioannidis 2020).

### Tradeoffs in Covid-19 research

Tradeoffs also exist in the choice of pandemic research methodologies, both in terms of the type of model and the data it is estimated on. For example, models may be estimated on data at the national or subnational level. Working at the level of the latter has the benefit of better controlling for country infection dynamics and avoiding possible issues arising from differences in pandemic accounting standards across countries (May 2020). Since data at the national level is broader in terms of country coverage than sub-national data, analyses at the level of the former may carry more external validity and will be less likely to fall prey to overfitting due to small sample noise. Finally, the tradeoff between aggregated (assuming homogeneity or pooled) or disaggregated (individual) modeling depends strongly on the amount and quality of the data.

The impact of policy interventions can be examined under the prism of three types of different models with their own set of tradeoffs: a) detailed models of epidemiological processes such as SIR-based models (Visscher 2020; Dehning et al. 2020; Davies et al. 2020), b) agent based modeling of transmission in a population (e.g., Aleta et al. 2020) and c) reduced form models that abstract away from these epidemiological processes and agent interactions at the micro-level, by using simple econometric models based on empirical data, rather than inference of complex epidemiological parameters. Epidemiological models, while desirable as they directly model the underlying mechanisms, are prone to overfitting in the presence of scant or low-quality, noisy data, leading to non-robustness (see Chin et al. 2020; Soltesz et al. 2020) since they require estimates of key parameters that are highly uncertain, and whose impact may reverberate significantly in highly non-linear, exponential growth models. No single approach dominates the rest, especially at the current stage of our understanding of the pandemic. Different approaches must be simultaneously pursued whilst acknowledging the limitations and advantages of each in an attempt to consolidate the findings.

As the literature on Covid-19 is burgeoning, I briefly survey the most similar studies, primarily those employing reduced-form equations. Early discourse was heavily influenced by the epidemiological modeling of the Imperial College COVID-19 Response Team (Flaxman et al. 2020), which concluded that lockdowns were a crucial and necessary strategy for the subduement of the pandemic. Sub-national data from six countries revealed that while not all specific NPIs have a significant impact, overall the implementation of NPIs significantly reduced infections (Hsiang et al. 2020), without however, explicitly accounting for how restrictive individual NPIs were. Dichotomizing the range of NPIs into less-restrictive (e.g., social distancing, discouragement of international and domestic travel, and large gatherings bans) and more-restrictive (e.g., stay-at-home and business closure orders) reveals no evidence of additional gains associated with the latter in ten countries at the subnational level (Bendavid et al. 2021). Similarly, stringent lockdowns in 24 European countries over the first half of 2020 did not significantly improve pandemic dynamics over less stringent lockdowns (Bjørnskov 2021). This was confirmed using data from 108 countries until May, 2020, (Bonardi et al. 2020) with the implication that NPIs may have a signalling effect leading to voluntary behavioral changes by citizens. Less stringent NPIs already provide a strong enough signal, so that there is nothing to gain from more stringent NPIs. The importance of voluntary behavioral changes is exemplified by the significant changes in mobility found to occur earlier than mandatory restrictions on movement (Abouk and Heydari 2021; Farboodi, Jarosch and Shimer 2020), and by the fact that mobility did not return to previous levels after the easing of lockdown restrictions (Gapen et al. 2020; Allcott et al. 2020).

I seek to combine the advantages of reduced-form approaches, whilst alleviating some of the drawbacks in their implementations to date. Specifically, earlier studies: a) eschewed behavioral components and agents’ incentives, b) estimated a single regression disavowing variable endogeneity, and c) were estimated on datasets from the first few months of the pandemic without the benefit of the currently accumulated data covering other developments such as newer variants.

### The importance of behavioral models

Models can be classified according to whether they are behavioral or not, i.e., are the behavioral responses of citizens and governments allowed to vary endogenously or are they assumed to be fixed? Agent-based models are by construct behavioral, however standard SIR models are not, and reduced-form work typically estimates single-equation regressions of effects of various NPI variables on either the confirmed case (and or death) growth rate or mobility data, thereby implicitly assuming exogeneity of these variables. However, it reasonable to believe that mobility is dependent on the severity of the NPIs in place, i.e., it may mediate the impact of NPIs on Covid-19 dynamics. Furthermore, NPI stringency may also be endogenous if governments base their policy decisions on recent epidemiological data and trends, e.g., infection growth. Consequently beyond modeling only SARS-CoV-2 dynamics, agents’ adaptive reactions to the situation merit attention: those of citizens (Cowling et al. 2020) (through mobility, precautionary and voluntary measures and other behavioral responses) and those of governments through the choice and timing of policies.

Beyond the foundational use of behavior in agent-based models, behavioral components can be introduced both to epidemiological and reduced form models. The former is significantly more common than the latter, which I will pursue in this study. The incorporation of behavioral components in SIR models leads to very different long-run predictions than models based on standard non-behavioral SIRs (Kermack and McKendrick 1927). A common finding is that the system tends to an equilibrium reproduction rate of 1 (Gans 2020), sometimes with oscillations if behavioral components operate with lags (Cochrane 2020; Atkeson 2021). These studies highlight the importance of modeling the evolution of behavioral responses, as they can lead to important qualitative, not just quantitative, changes in pandemic dynamics. The canonical behavioral response depends on the perceived risk of infection and severity; e.g., consumers’ behavioral changes in shopping habits were predominantly of their own volition rather than through the imposition of legal restrictions (Goolsbee and Syverson 2021). Behavioral incentives can have a stabilizing effect on the infection growth rate, as the higher it is, the more likely citizens are to react by adjusting their behavior to decrease the chances of infection. However, it is important to also bear in mind that individual behavior may become more reckless as risks are mitigated—see the risk-compensation literature (Peltzman 1975). For example, as the pandemic starts to wane, citizens may adopt more risky practices, thereby slowing down the decrease in the infection rate. Similarly, extensive testing and vaccinations may elicit adverse behavioral responses if citizens believe they are less likely to contract Covid-19 and infect others.

### The importance of beliefs and expectations

Importantly, there may be latent (unobservable) variables, such as culture (Laliotis and Minos 2020), at the country-level that jointly influence key variables that are typically assumed to be exogenously determined instead. For example, low social capital and trust in government may affect case growth rates directly and indirectly, through other explanatory variables. Suppose that low-trust countries also tend to have inadequate public health systems. This will have a direct impact on case and mortality growth due to inadequate health care, but also an indirect effect. The government, knowing that the health system is weak (e.g., scarce intensive care units), may impose more stringent NPIs compared to countries with better health systems, in a attempt to prevent them from filling to capacity. Expectations and beliefs of citizens and governments can further introduce endogeneity. Low trust between citizens and government institutions may lead citizens to underestimate the severity of the situation as presented by governments, and the government, expecting this, may act to impose stricter restrictions in anticipation of the weaker behavioral response by its citizens.

### Simultaneously modeling endogeneity and unobservable variables, behavioral incentives and beliefs

Acknowledging that confirmed case and death growth, mobility and government policies are endogenous requires a system of four equations to fully model these interactions and avoid biases resulting from simpler regressions that implicitly impose exogeneity. I complement the literature by employing econometric procedures, namely structural multi-equation modeling including unobservable or latent variables and their effects, to improve upon the external validity of prior analyses. Such a system of equations can be viewed through the lens of game theory, that is, the modeling of different agents, who react to each others strategies and expectations thereof. I propose, an admittedly rudimentary, model of the adaptive interactions of these three agents.^1^ My approach complements existing studies by investigating a large number of countries, with the associated benefits of pooling estimates for robustness, but other disadvantages such as the assumption of homogeneity across countries and the need to focus on aggregated measures of key endogenous variables–rather than their individual components (Haug et al. 2020)–to avoid a computationally-intractable and unidentifiable system of equations. A central question is whether there are decreasing returns to the effectiveness of NPIs with severity, and if so, to estimate the approximately optimal severity of government policies. This remains a critical question not only for future pandemics, but also because NPIs may be valuable even during the vaccination phase (Rella et al. 2021).

## Methods

National-level data from 132 countries covering the time period from 15th February 2020 to 14th April 2021 was compiled, extending previous analyses to data including the appearance and spread of the B.1.1.7 and B.1.351 variants detected late 2020, and the more recently discovered variants such as P.1. Confirmed case and death counts, vaccinations, and tests, were download from the Covid-19 Data Hub (Guidotti and Ardia 2020), which also included the implementation of NPIs–a composite score or Stringency Index–sourced originally from the Oxford COVID-19 Government Response Tracker (Hale et al. 2021). Mobility data from the Google Community Mobility Report (Google 2020) was merged with the datafile from the aforementioned database.

The growth in confirmed cases (deaths) was calculated as the log difference in the cumulative confirmed cases (deaths) for two consecutive days multiplied by 100 (i.e., they can be interpreted as approximate percentage growth rates). The summary statistics for growth in confirmed cases are: # of obs. 53,279, mean=2.74%, stdev.=8.87% and for deaths: # of obs. 49,366, mean=1.997%, stdev.=6.84%. To allow for a non-linear relationship between the Stringency Index and case/death growth, a semi-parametric approach was implemented by subdividing the SI (ranging continuously from 0 to 100) into a baseline of no restrictions (0) and the deciles (1–10, 11–20, …, 91–100). The set of (non-restrictive) policies that we examine are: the testing policy (TP) variable, which takes on three levels (*l*) of 0 (no testing) through to 3 (open testing), contact tracing (CT) policy takes on levels of 0 through 2, the proportion of the population tested per day (TPop) and the cumulative percentage of the number of vaccinations compared to the country’s population (V)—this can be greater than 100% due to some vaccines requiring more than one dose.

I address the issues identified above by simultaneously estimating four generalized structural equations to model the complex inter-relationships between variables—see Figure 2 for a graphical representation of the causal structure and equations 1–4. The dependent variables for each equation are: (eq. 1) the growth rate of confirmed cases (*Ċ*), (eq. 2) the growth rate of confirmed deaths 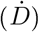, (eq. 3) the seven-day moving-average of mobility *M*_*i,t*_ and (eq. 4) the ordered categorical variable 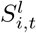 derived from the stringency index.

**Figure 2:**
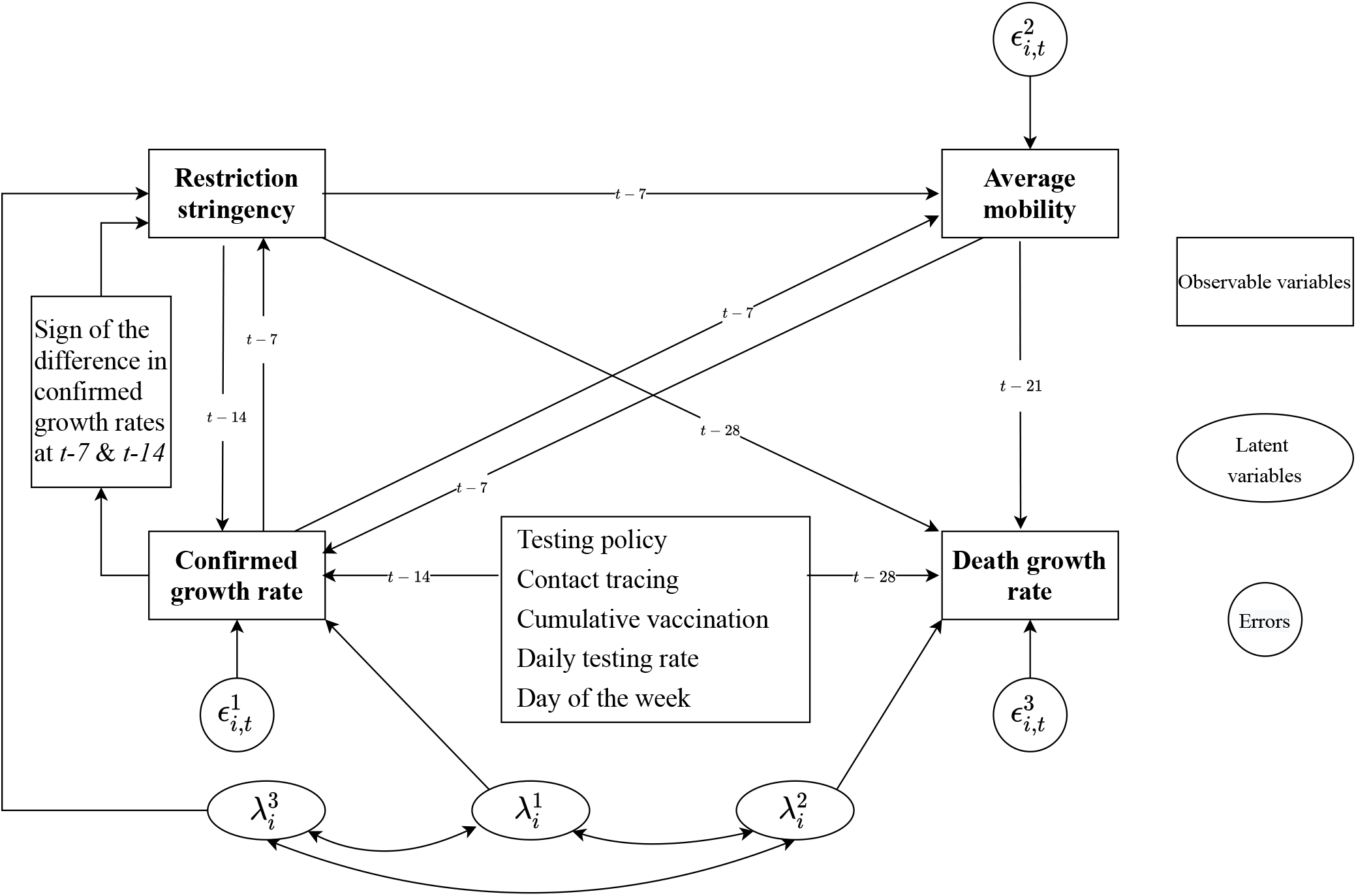
A 4-equation structural model of Covid-19 dynamics

Equations 1–4 below completely define the econometric model. It incorporates lagged relationships between the case (and death) growth rates, and other variables arising from the approximate delay in symptom onset and case confirmation and the timing of exposure to the virus (Rothan and Byrareddy 2020). The lags for the growth rate in deaths are longer than those for cases (by 14 days) to reflect the fact that on average deaths occur at a much later time after infection, e.g., due to deaths occurring after lengthy hospitalization in intensive care units. The exact determination of the lags is not critical, as there is a high degree of autocorrelation of the key lagged variables. To simplify the model, all lags and moving-averages were multiples of 7 days to remove the effects of systematic daily variations.

The 7-day moving average of the percentage change in mobility (compared to the pre-Covid baseline for each country) is denoted by *M*_*i,t*_—this was constructed by an equal-weighting of three individual variables separately measuring mobility towards groceries and pharmacies, transit stations and workplaces. Since this variable is normalized at a different baseline per country, additional random effects at the country level were not implemented in equation 3, in contrast to all other equations.

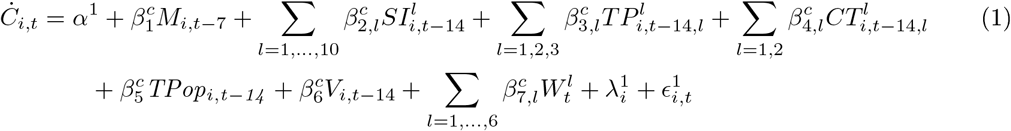

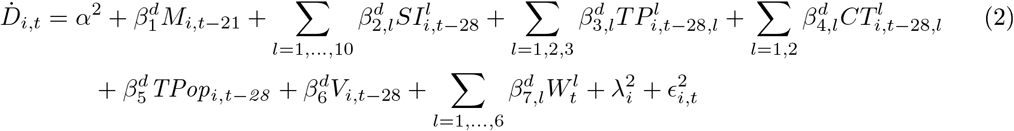

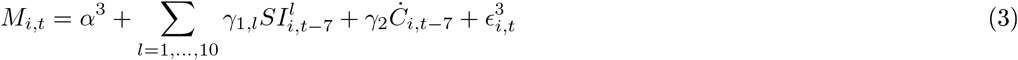

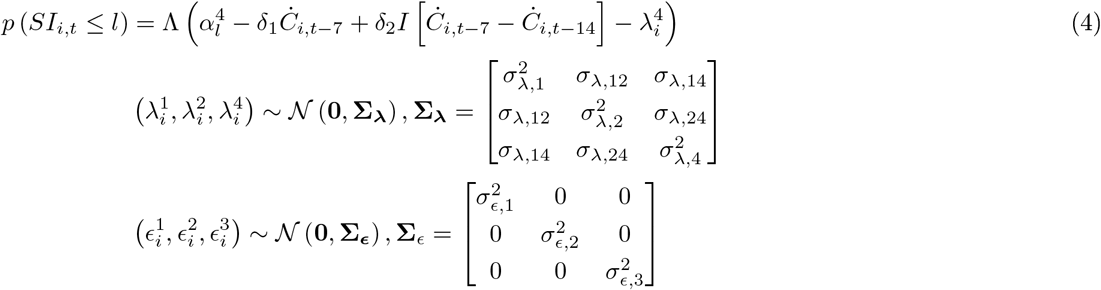

In eq. 4, 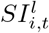 is an ordered variable of the stringency index ranging in levels from 0 to 10, in equation 1 it enters as a set of dummy variables. An indicator function captures government hysteresis or path-dependence *I* [*Ċ*_*i,t*−7_ *− Ċ*_*i,t*−14_], which is equal to 1 if the case growth rate has been increasing or zero if it has been decreasing. If the estimated coefficient *δ*_2_ *>* 0 then this means that for the same level of case growth, governments will on average be more likely to impose more stringent NPIs if the case growth trend is negative than if it were positive.

The initial date used in the estimation varied by country as it was to set to the first day with a confirmed case. Any missing datapoints for the confirmed cases and deaths, number of tests and vaccinations at time *t* were set equal to the value at time *t −* 1; note, this was done on the raw data of these variables, which were coded as cumulative sums until time *t*. Country-level unobservable characteristics were modeled using unique random effects 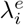 for equations *e* =1, 2 and 4, allowing for covariance between all the random effects to capture the joint impact of unobservable country characteristics. Fixed-effects for the day of week are denoted by 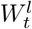.

Possible hysteresis, or path-dependence, in governments’ determination of the level of NPI stringency is also modeled, conditional on whether the case growth rate has recently been declining or increasing. The hypothesis is that governments are slower at scaling back stringent NPIs as the pandemic threat recedes than they are in implementing them during outbreaks. Fixed-effects accounted for the possible influence of the day of the week on case and death rates. Cluster-robust standard errors were employed to account for heteroskedasticity and within-panel dependence of errors. Of course, the usual disclaimers hold regarding the use of observational data and the causal assumptions embedded in the chosen structural equations and variable relationships.

## Results

Detailed regression results can be found in Tables 2 and 3, below I present the main estimates graphically.

**Table 1:**
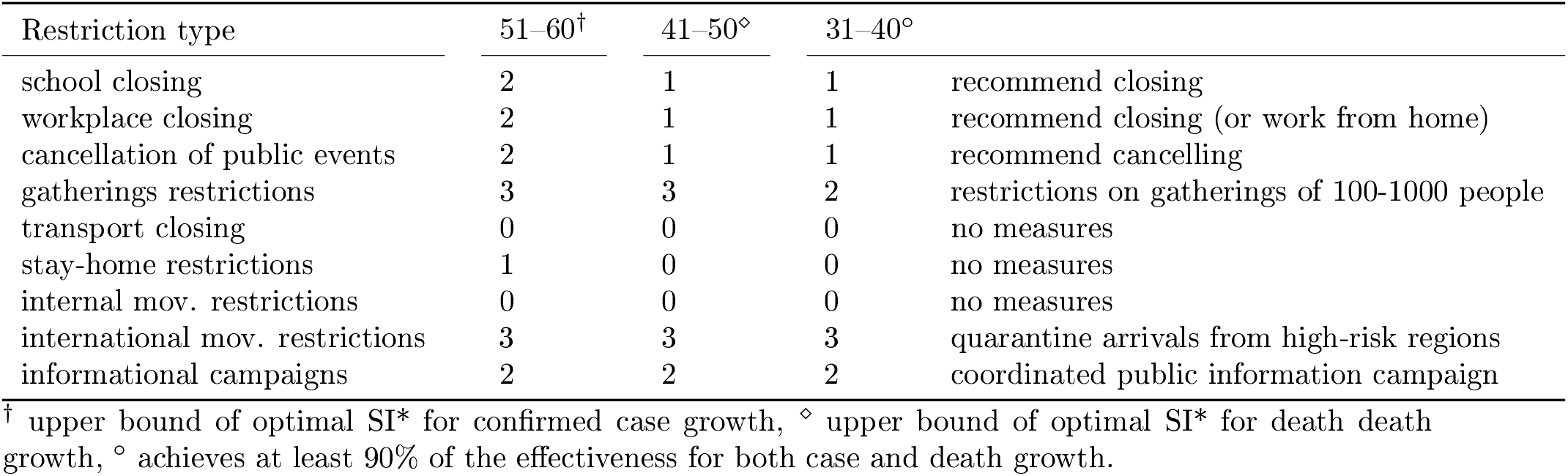
Stringency index levels and their (median) constitutent NPI levels

**Table 2:** Generalized structural equation model of *Ċ*_*i,t*_, 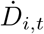, *M*_*i,t*_ and *SI*_*i,t*_

| $\dot{C}_{i,t}$ | | $\dot{D}_{i,t}$ | | $M_{i,t}$ | | $SI_{i,t}$ | |
| --- | --- | --- | --- | --- | --- | --- | --- |
| $M_{i,t-7}$ | -0.0417***<br>[-0.0578,-0.0256] | $M_{i,t-21}$ | -0.0162*<br>[-0.0304,-0.00199] | | | | |
| $SI_{i,t-14}$ | | $SI_{i,t-28}$ | | $SI_{i,t-7}$ | | | |
| 1-10 | -4.522<br>[-10.40,1.350] |  | -7.336***<br>[-10.97,-3.701] | 1-10 | -3.346<br>[-7.675,0.983] |  |  |
| 11-20 | -5.681*<br>[-10.01,-1.352] |  | -7.938***<br>[-10.85,-5.022] | 11-20 | -3.264<br>[-10.44,3.910] |  |  |
| 21-30 | -10.44***<br>[-14.73,-6.153] |  | -12.45***<br>[-15.16,-9.734] | 21-30 | -6.421*<br>[-12.29,-0.550] |  |  |
| 31-40 | -13.45***<br>[-17.42,-9.487] |  | -14.96***<br>[-17.71,-12.22] | 31-40 | -12.21***<br>[-15.98,-8.438] |  |  |
| 41-50 | -14.13***<br>[-18.04,-10.22] |  | -15.46***<br>[-18.14,-12.79] | 41-50 | -13.90***<br>[-17.54,-10.26] |  |  |
| 51-60 | -14.81***<br>[-18.70,-10.92] |  | -15.80***<br>[-18.45,-13.14] | 51-60 | -15.68***<br>[-19.40,-11.96] |  |  |
| 61-70 | -15.25***<br>[-19.18,-11.33] |  | -16.10***<br>[-18.80,-13.40] | 61-70 | -19.84***<br>[-23.80,-15.88] |  |  |
| 71-80 | -15.28***<br>[-19.19,-11.36] |  | -15.91***<br>[-18.59,-13.22] | 71-80 | -28.68***<br>[-32.76,-24.59] |  |  |
| 81-90 | -15.10***<br>[-19.04,-11.17] |  | -15.83***<br>[-18.57,-13.10] | 81-90 | -34.90***<br>[-39.67,-30.13] |  |  |
| 91-100 | -14.19***<br>[-18.23,-10.15] |  | -15.07***<br>[-17.84,-12.30] | 91-100 | -53.46***<br>[-59.67,-47.25] |  |  |
| $Tpop_{i,t-14}$ | -0.184<br>[-0.404,0.0355] | $Tpop_{i,t-28}$ | -0.126<br>[-0.315,0.0629] | $\dot{C}_{i,t-7}$ | -0.214***<br>[-0.255,-0.174] | $\dot{C}_{i,t-7}$ | 0.0484***<br>[0.0354,0.0613] |
| $V_{i,t-14}$ | -0.0170*<br>[-0.0316,-0.00242] | $V_{i,t-28}$ | -0.0187*<br>[-0.0352,-0.00230] | | | $I \left[ \begin{array}{l} \dot{C}_{i,t-7} \\ -\dot{C}_{i,t-14} \end{array} \right]$ | 0.531***<br>[0.453,0.609] |
| $TP_{i,t-14}(1)$ | -4.385***<br>[-6.824,-1.946] | $TP_{i,t-28}(1)$ | -2.603*<br>[-4.671,-0.535] | | | | |
| $TP_{i,t-14}(2)$ | -6.474***<br>[-8.964,-3.983] | $TP_{i,t-28}(2)$ | -4.247***<br>[-6.369,-2.125] | | | | |
| $TP_{i,t-14}(3)$ | -7.390***<br>[-9.926,-4.854] | $TP_{i,t-28}(3)$ | -4.875***<br>[-7.086,-2.664] | | | | |
| $CT_{i,t-14}(1)$ | -0.77<br>[-1.973,0.434] | $CT_{i,t-28}(1)$ | -0.567<br>[-1.584,0.450] | | | | |
| $CT_{i,t-14}(2)$ | -0.82<br>[-1.986,0.345] | $CT_{i,t-28}(2)$ | -0.572<br>[-1.586,0.442] | | | | |
| $\alpha^1$ | 21.96***<br>[17.92,25.99] | $\alpha^2$ | 20.71***<br>[18.30,23.13] | $\alpha^3$ | 3.328*<br>[0.0958,6.560] | | |
| $\sigma_{\lambda,1}^2$ | 4.299***<br>[3.312,5.287] | $\sigma_{\lambda,2}^2$ | 2.870***<br>[2.076,3.665] | | | $\sigma_{\lambda,4}^2$ | 3.240***<br>[2.350,4.130] |
| $\sigma_{\epsilon,1}^2$ | 22.22***<br>[17.82,26.62] | $\sigma_{\epsilon,2}^2$ | 33.72***<br>[29.39,38.04] | | | $\sigma_{\epsilon,4}^2$ | 200.4***<br>[160.2,240.5] |
| $\sigma_{\lambda,12}=3.429***$ [2.683,4.176] , $\sigma_{\lambda,14}=2.478***$ [1.798,3.158] , $\sigma_{\lambda,24}=2.070***$ [1.479,2.661] | | | | | | | |
| [95% CI],* p<0.05, **p<0.01, *** p<0.001 |  |  |  |  |  |  |  |

**Table 3:** Other estimates from the structural equation model of *Ċ*_*i,t*_, 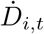 *M*_*i,t*_ and *SI*_*i,t*_

| $\dot{C}_{i,t}$ | | $\dot{D}_{i,t}$ | $SI_{i,t}$ | |
| --- | --- | --- | --- | --- |
| Monday | -0.0198<br>[-0.167,0.128] | 0.153<br>[-0.0432,0.349] | $\alpha_1^3$ | -8.621***<br>[-9.903,-7.340] |
| Tuesday | 0.14<br>[-0.0678,0.347] | 0.189<br>[-0.0111,0.389] | $\alpha_2^3$ | -6.679***<br>[-7.437,-5.920] |
| Wednesday | 0.332***<br>[0.173,0.491] | 0.238*<br>[0.0540,0.423] | $\alpha_3^3$ | -4.625***<br>[-5.252,-3.999] |
| Thursday | 0.278***<br>[0.145,0.412] | 0.210*<br>[0.00722,0.413] | $\alpha_4^3$ | -3.281***<br>[-3.778,-2.784] |
| Friday | 0.392***<br>[0.263,0.521] | 0.18<br>[-0.000637,0.361] | $\alpha_5^3$ | -1.993***<br>[-2.413,-1.574] |
| Saturday | 0.103<br>[-0.00538,0.211] | 0.123<br>[-0.0604,0.307] | $\alpha_6^3$ | -0.808***<br>[-1.192,-0.425] |
| | | | $\alpha_7^3$ | 0.232<br>[-0.131,0.594] |
| | | | $\alpha_8^3$ | 1.289***<br>[0.944,1.635] |
| | | | $\alpha_9^3$ | 2.617***<br>[2.249,2.984] |
| | | | $\alpha_{10}^3$ | 4.470***<br>[4.016,4.924] |

### Highly correlated latent (unobservable variables) influence both government policy and confirmed case/death growth rates

The estimated covariances (and associated correlation) between the country-level latent variables in equations 1, 2 and 4 are all positive and significantly different from zero at the 0.001% level: *ρ*_*λ*,12_=0.98 [95% CI: 0.95,1.00], *ρ*_*λ*,14_=0.66 [0.54,0.79], *ρ*_*λ*,24_=0.68 [0.56,0.80]. This validates our hypothesis that there exist significant unobservable variables that may simultaneously influence case and death rates, but also government responses to the pandemic in terms of the severity of restrictions. Ignoring these relationships by estimating a single regression of case or death growth rates, as is commonly done in the literature, would have led to biased estimates.

### Adaptive expectations of the risk of infection impact non-residential mobility

Beyond the effect of NPIs, citizens react to increases in the 7-day lag in the growth rates of confirmed cases by reducing mobility. This is consistent with a theory of citizens forming adaptive expectations about the severity of the pandemic at any point in time and the risk of contraction based on the data. The impact of the daily growth in confirmed cases is 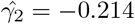[-0.255,-0.174]. Putting the effect size into perspective, an increase in the case rate of 1% point, would result on average in a relatively weak 0.21% point reduction in mobility compared to the baseline.

### The indirect links between NPIs and confirmed cases/deaths

The indirect links consist of two components in each case, (*SI → M*) and (*M → Ċ*), (*SI → M*) and 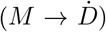. The following results will establish that both indirect components are significantly different from zero. Consequently, non-residential mobility acts as the mediator in this relationship between NPI stringency and case/death growth rates. First I report the evidence regarding the *SI → M* component that is common to both instances and then the second component.

### Higher NPI severity restricts non-residential mobility

Non-residential mobility is clearly impacted by the SI (see the estimates in Table 2 and Figure 3 below), as the null hypothesis that all SI dummy variables are equal to zero is rejected (*χ*^2^ (10) = 428.38, *p <* 0.0001). Furthermore, it is monotonically decreasing in the ten SI ranges.

**Figure 3:**
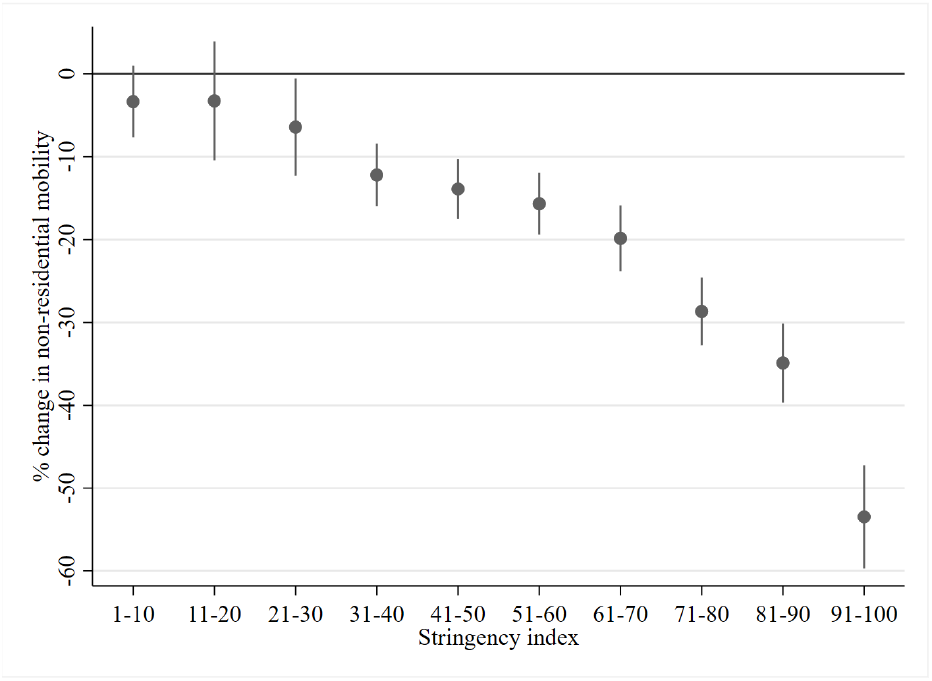
Impact of policy Stringency index on the percentage change in non-residential mobility (*γ*_1,*l*_)

### Lower non-residential mobility increases case and death growth rates

The theoretical motivation behind restricting mobility through lockdown measures is that its reduction will lead to a fall in the number of social interactions, with the hope that this will reduce transmission and infection. However, a reduction in non-residential mobility must, by definition, be mirrored by an increase in the time spent within residences—indeed, they are highly negatively correlated in the Google mobility data; the median correlation within panels is -0.89. Consequently, reductions in non-residential mobility may have a detrimental effect on case/death growth as although transmission outside the home may be reduced, within-household transmission may be enhanced as people spend increasingly more time within the confines of a closed space with others (Sun et al. 2021). Since the two effects work in opposite directions, whether restricting non-residential mobility reduces case/death growth or not must be resolved empirically. Our finding that the coefficient of *M*_*i,t*−7_ is significantly negative for cases, 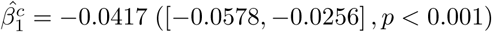 and similarly *M*_*i,t*−21_ for deaths 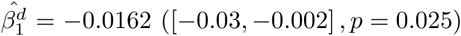, supports the hypothesis that the benefits of reduced non-residential mobility are more than outweighed by the detrimental effects of increased within-household transmission (conditioning on the stringency of government policies). The effect size is moderate, as a 10% point decrease in mobility leads to an increase in 0.4% points of the case growth rate.

### The direct link between SI and case/death growth rates

Turning to the direct effect of the stringency index of restrictions on case growth rates, the hypothesis that the set of 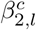 estimates are all equal to zero is rejected (*χ*^2^ (10) = 137.28, *p <* 0.0001). Furthermore, there is evidence of a nonlinear relationship, with strongly decreasing returns to the effectiveness of restrictions with increasing severity (see Table 2, and note that the conclusions are similar for death growth rates).

### The total effect of NPI stringency on case/death growths and the optimal level of SI

The total effect of NPI stringency on case growth can be computed by adding the direct *β*_2,*l*_ and indirect paths *γ*_1,*l*_ *× β*_1_ for each stringency level of *l*. This combined effect of SI on *Ċ*_*i,t*_ and 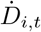 is documented in Table 4 and presented graphically below in Figure 4.

**Table 4:** Total effects of SI ranges on the growth rate of confirmed cases

| | Coef. | $p$ | 95% CI | |
| --- | --- | --- | --- | --- |
| 1–10 | -4.383 | 0.140 | -10.202 | 1.436 |
| 11–20 | -5.544 | 0.012 | -9.880 | -1.209 |
| 21–30 | -10.173 | 0.000 | -14.473 | -5.874 |
| 31–40 | -12.945 | 0.000 | -16.883 | -9.007 |
| 41–50 | -13.549 | 0.000 | -17.424 | -9.674 |
| 51–60 | -14.156 | 0.000 | -18.013 | -10.298 |
| 61–70 | -14.426 | 0.000 | -18.319 | -10.534 |
| 71–80 | -14.080 | 0.000 | -17.953 | -10.208 |
| 81–90 | -13.647 | 0.000 | -17.531 | -9.763 |
| 91–100 | -11.958 | 0.000 | -15.906 | -8.010 |

**Figure 4:**
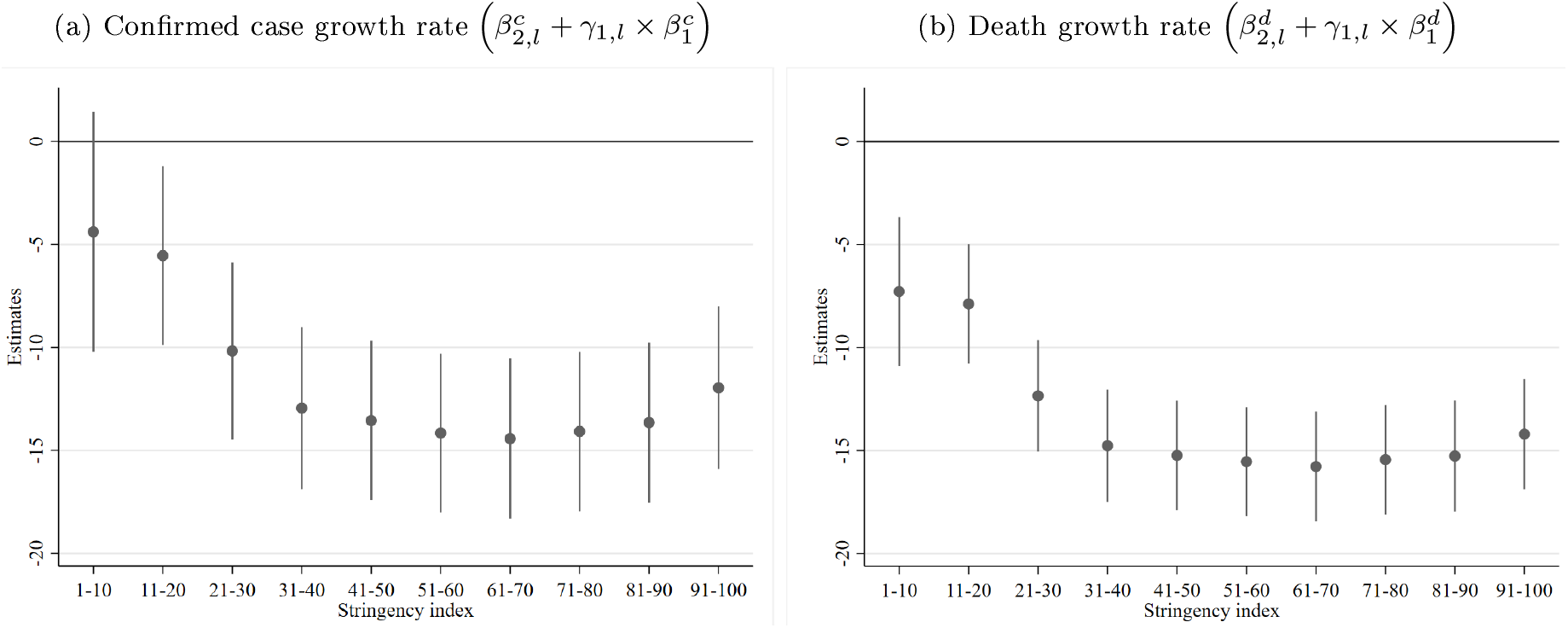
Total effect of the level of the Stringency Index

The maximum impact of SI on case growth is observed for the SI range of 61–70. We test the difference in effectiveness of all other levels against the most effective range 61–70, correcting for multiple comparisons using the Sidak correction.^2^ There is no difference in effectiveness for the ranges 51–60 and 71–80 (see Table 5 for the test statistics); the ranges (81–90, 91–100) are significantly less effective than the range of 61–70. Consequently, there are no further gains to be achieved beyond the SI range of 51–60. The socially optimal SI range, however, must account not only for the positive effects of NPIs, but also for the significant economic and other health-related costs that result from restrictions (for example, see Miles, Stedman and Heald 2020; Susskind and Vines 2020; Altman 2020). While this would require a full cost-benefit analysis (Rowthorn and Maciejowski 2020;Appleby 2020) that is beyond the scope of this paper, we can derive the approximate upper bound of the socially optimal level of the SI with a single assumption about the cost profile of different SI levels. Namely, I only assume that the costs are monotonically increasing in the SI level. Consequently, without the need to quantify costs, I conclude that the upper bound of the socially optimal SI level, SI* is 51–60, i.e., the minimum SI range that is not significantly different from the maximum effect at 61–70.

**Table 5:** Difference between the maximum effectiveness level of restrictions for confirmed cases (61–70)

| Relative to 61–70 | $\chi^2(1)$ | $p$ |
| --- | --- | --- |
| 1–10 | 18.310 | 0.0002 |
| 11–20 | 43.220 | 0.0000 |
| 21–30 | 21.300 | 0.0000 |
| 31–40 | 13.910 | 0.0017 |
| 41–50 | 9.410 | 0.0193 |
| 51–60 | 2.550 | 0.6496 |
| 71–80 | 3.480 | 0.4392 |
| 81–90 | 8.340 | 0.0343 |
| 91–100 | 43.400 | 0.0000 |
| Joint | 101.710 | 0.0000 |

While the quantification of the exact costs is an important endeavour, it is fraught with many difficulties, such as converting non-economic outcomes into monetary terms and would require many assumptions about highly uncertain possible effects, many in the distant future. By contrast, this upper-bound on SI* is extremely robust, albeit less informative. The above derivation of SI* is based on statistical significance, however it is possible that lower SIs may be statistically significantly different from SI*, but that the difference in the effect size is practically of little importance. Indeed, the ratio of the effect sizes of an SI range of 31–40 relative to the average of the 51–80 range, is 91% [85%, 97%]. The relative effectiveness of the even laxer 21–30 range is 72% [57%, 86%]. If the costs of NPIs increase quickly with NPI stringency, it is conceivable that moderately severe policy responses in the 30–40 SI range (and possibly, even 21–30 SI) may in fact be close to the socially optimal SI* (arising from a full cost-benefit analysis), as it already achieves 91% effectiveness without accounting for costs.

Similarly, the maximum effectiveness on death growth rates is also observed for an SI range of 61–70. We test the difference in effectiveness of all other levels against this range. There is no significant increase in effectiveness for levels beyond 41–50 (see Table 7). In terms of the effect size relative to the average of the 41–90 range, an SI range of 31–40 achieves 93% [87%, 98%] of the effectiveness of the former and the laxer 21–30 range achieves 73% [58%, 88%].

**Table 6:**
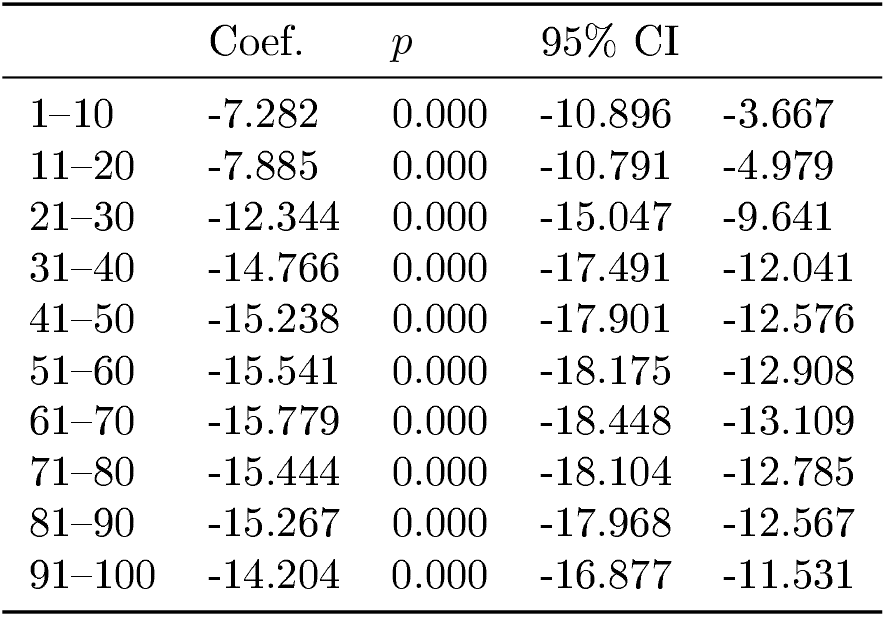
Total effects of SI ranges on the growth rate of deaths

| | Coef. | $p$ | 95% CI | |
| --- | --- | --- | --- | --- |
| 1–10 | -7.282 | 0.000 | -10.896 | -3.667 |
| 11–20 | -7.885 | 0.000 | -10.791 | -4.979 |
| 21–30 | -12.344 | 0.000 | -15.047 | -9.641 |
| 31–40 | -14.766 | 0.000 | -17.491 | -12.041 |
| 41–50 | -15.238 | 0.000 | -17.901 | -12.576 |
| 51–60 | -15.541 | 0.000 | -18.175 | -12.908 |
| 61–70 | -15.779 | 0.000 | -18.448 | -13.109 |
| 71–80 | -15.444 | 0.000 | -18.104 | -12.785 |
| 81–90 | -15.267 | 0.000 | -17.968 | -12.567 |
| 91–100 | -14.204 | 0.000 | -16.877 | -11.531 |

**Table 7:**
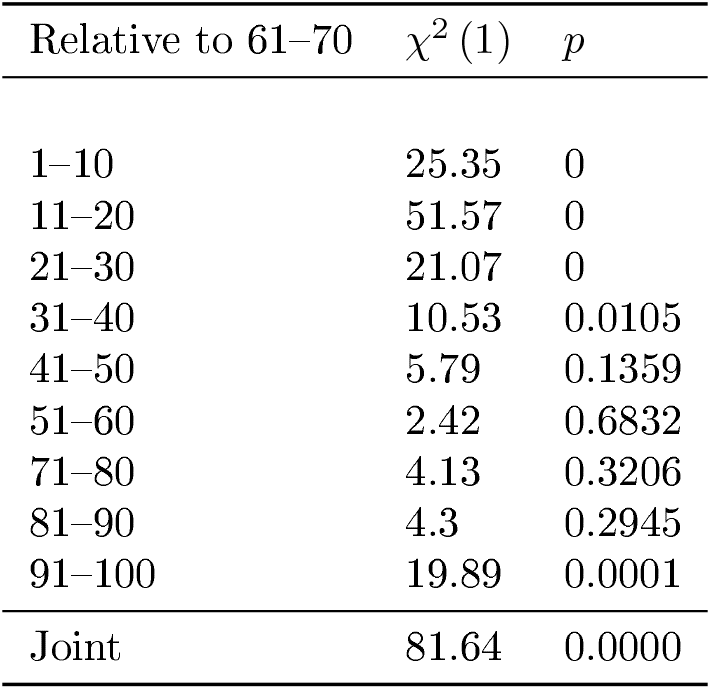
Difference between the maximum effectiveness level of restrictions for deaths (61–70)

Finally, while the SI index aggregates various individual NPI policies, examining the median values of the individual policies in the dataset for each SI range can be informative—see Table 1. Note, that for the 31–40 SI range, which achieves at least 90% of the maximum impact, the median policies do not include any level of restriction on transport and internal movement, and no stay-home restrictions. Furthermore, only recommendations for closing schools and workplaces and cancellation of public events were issued, i.e., these were not mandatory. The only stringent individual policies typically arising in the 31–40 SI range were quarantining high-risk cases from international travel and restrictions on gatherings of 100-1000 people; however, these two policies are not reflective of citizens’ everyday behavior. Consequently, voluntary–rather than mandatory–behavioral changes are more important drivers of the impact of NPIs on case (and death growth).^3^ This is consistent with other studies that also conclude that the flattening of NPI effectiveness with increasing stringency reflects a relatively stronger voluntary rather than mandatory component to behavioral changes (e.g., Allcott et al. 2020; Bjørnskov 2021; Bonardi et al. 2020; Bendavid et al. 2021).

Whence arises this voluntary behavioral change? As we showed above, part of it is from citizens’ expectations of the risk of infection and severity as captured by 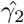 in equation 3, whose effect size, however, was found to be relatively small. The rest, and majority of the voluntary behavioral change, is likely due to the signalling value of policy decisions, i.e., citizens can also use information about the stringency of the government measures to infer the severity of the pandemic. This implies that recommendations by governments, for SI ranges of up to 40, were heeded by citizens, who significantly changed their behavior in ways beyond those captured by mobility in eq. 3, e.g., preventative measures such as diligent hand washing and mask-wearing, effective self-isolation when infected et cetera.

### Extensive public testing significantly reduces case and death growth, contact tracing does not

Figure 5 presents the estimated coefficients and associated 95% confidence intervals—see Table 2 for detailed regression results. All three levels of the testing regime are jointly significantly different from zero (*χ*^2^ (3) = 66.03, *p <* 0.0001), leading to progressively greater declines in case growth as testing becomes more extensive (robust to multiple comparison Sidak corrections).^4^ Note, that the most extensive testing policy has an impact of -7.39 [-9.926,-4.854] which is 51% [27%,76%] of that of the most impactful SI range (61–70).

**Figure 5:**
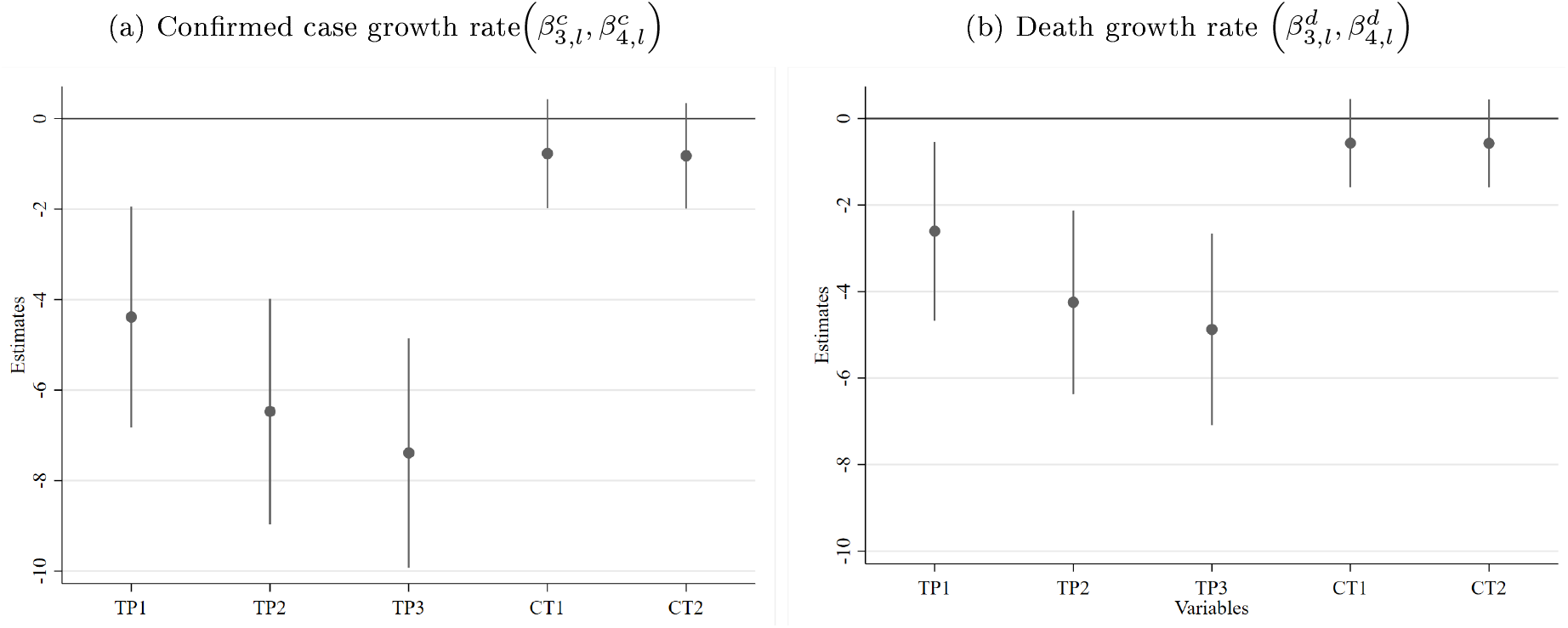
Estimated coefficients across levels of testing policy and contact tracing

Similarly, testing policy significantly reduces the death growth rate (*χ*^2^ (3) = 42.81, *p <* 0.0001). Apart from the monetary costs associated with implementing a testing regime it does not impose other negative social externalities, in contrast to lockdowns. Coupled with the significant impact on Covid-19 dynamics, this renders extensive testing a desirable tool.

Contact tracing of any level does not have a significant impact on confirmed case growth (*χ*^2^ (2) = 1.93, *p* = 0.38) or death growth (*χ*^2^ (2) = 1.31, *p* = 0.52). One should keep in mind that contact tracing may still be effective if the number of daily new cases is small, when efficient tracing is more manageable. However, there remain important challenges to scaling contact tracing (Chowdhury et al. 2020; Quilty et al. 2021; Kretzschmar et al. 2020) that could hinder its effectiveness during significant outbreaks.

### The proportion of the population tested daily does not significantly affect cases and deaths

While both estimates are negative, as expected, increasing the proportion of daily tests does not significantly reduce the case growth, 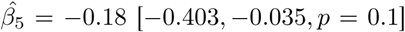, nor the growth rate in deaths, -0.126 [*−*0.315, 0.063, *p* = 0.191]. Note, that an earlier analysis with datapoints till the end of January 2021 only, revealed a statistically significant impact. The inclusion of different levels of the ordinal testing policy variable may partially capture this effect, as they will be correlated to some degree with the proportion tested, i.e., extensive testing as coded in the ordinal variable would likely imply more testing. Finally, this may be the result of implicitly assuming exogeneity, however testing may also be endogenously determined as governments are likely to step up testing during phases with higher transmission.

### Vaccination reduces case and death growth rates

Despite few datapoints where vaccination was already well underway, each 1% point increase in the cumulative vaccination % results in a reduction of the case growth of -0.017% points [*−*0.032, *−*0.002, *p* = 0.022] and in the death growth rate -0.0187% points [*−*0.035, *−*0.002, *p =* 0.026]. Note that the median (non-zero) cumulative vaccination % across countries is only 2.6% and the 10th and 90th percentiles are 0.07% and 19.8%, respectively. Consequently, these relatively low estimates should not be assumed to extrapolate for higher vaccination levels, especially since this is likely to be a nonlinear relationship in reality, i.e., the effect of vaccinations will increase at an accelerating rate as it approaches the herd immunity level.

### Government policy is endogenous and exhibits hysteresis

Government policy is strongly endogenous, in contrast to the common implicit assumption of exogeneity. The seven-day lagged confirmed growth rate coefficient 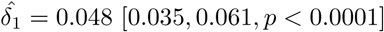 is positively related to NPI severity. Furthermore, I find significant hysteresis in the de-escalation of NPIs, that is for the same case growth, NPIs are significantly more stringent if the case growth has been falling recently, than the opposite: 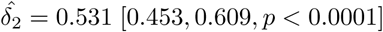.

## Discussion

A four-equation structural model of multiple agents (SARS-CoV-2 virus, citizens and government) capturing the basic dynamics of their endogenous evolution revealed the following. Recall that the Stringency Index of NPIs ranges from 0 (no measures taken) to a maximum of 100. For confirmed case growth rates, there were no significant gains to be had beyond an SI range of 51–60 moreover, 91% of this effect size can be achieved with an SI range of 31–40. For death growth rates, no significant gains were to be achieved beyond an SI range of 41–50 and 93% of this effect size can be accomplished with an SI range of 31–40. An open testing policy has approximately half the benefits of the optimal NPIs without incurring the societal costs associated with long-term restrictions. Furthermore, the finding that decreases in non-residential mobility, and therefore increases in its complement, time spent at residences, increase the growth rate of confirmed cases and deaths is aligned with contact tracing analyses of heightened transmission risk within a household, compared to the wider community (Sun et al. 2021) and earlier work concluding that shelter-in-place orders did not reduce Covid-19 infection and mortality rates (Berry et al. 2021).

### Interpretation and implications for policy

What implications does this have for policy? I report in Table 1 the median values of the individual NPI constituents of the SI index for these upper bounds on the socially optimal level of NPI stringency for both confirmed cases (51–60) and deaths (41–50), and the range of 31–40, which is near-optimal. Note, that there is significant heterogeneity across individual NPIs in terms of their severity. The most severe implemented restrictions are those on gatherings and international movement. At the other extreme, regarding transport and internal movements, no restrictions or recommendations were made in these three SI ranges. Stay-home restrictions and recommendations are also absent with the exception of a recommendation to stay home associated with the 51–60 range. Moderately severe restrictions are typically implemented for schools and workplaces, that is, the optimal upper bound for confirmed cases enforces partial closing (targeted, not across the board) whereas that for death rates and the near-optimal SI range (31–40) call only for recommendations to work from home.

These findings are generally aligned with studies finding that more severe restrictions were not significantly more effective than less restrictive policies (Bendavid et al. 2021; Haug et al. 2020; Bjørnskov 2021; Bonardi et al. 2020). However, they stand in contrast to others that concluded that strict lockdowns were effective (Flaxman et al. 2020; Islam et al. 2020), whilst differing in some policy interventions such as school and workplace closures, but agreeing on others such as mass gathering restrictions (Islam et al. 2020). Differences with these studies may arise due to their more limited time-series and/or the lack of explicit behavioral modeling of citizens’ reactions to the pandemic. The majority of the effect of NPIs on case and death growth rates could be attributed to voluntary behavioral changes rather than mandatory, government-imposed imperatives. Note also, that the 31–40 range typically includes a coordinated public informational campaign aimed at influencing voluntary behavior. This highlights the importance of modeling the behavioral incentives of both governments and citizens in conjunction with the pandemic dynamics.

Importantly, the study’s conclusions were shown to be robust to the rectification of econometric issues such as endogeneity and correlated unobservable latent variables, and accounting for citizens’ and governments’ voluntary behavior.

### Strengths and limitations of this study

The simultaneous modeling of pandemic dynamics with behavioral models of citizens’ behavioral adaptation to the pandemic, along with a model of governments’ decision-making processes regarding policy implementation is the primary strength of this study with respect to earlier work. Identification of this more sophisticated model with behavioral components was made possible by the larger amount of accumulated data including testing and vaccination rates. Nonetheless, certain simplifications were still necessary to ensure identification and to rein in the computational complexity of the estimation processes. These simplifications included examining the effects of nations’ stringency index compiled from individual NPIs, rather than examining each NPI separately. Similarly, an average measure of the change in mobility was used instead of disaggregated sub-measures of the type of mobility. Furthermore, while random-effects allowed for variation across countries in unobservable variables, estimates of the variables of interest (NPIs and other interventions) were pooled across countries.

### Unanswered questions and further research

As more data becomes available over time, future research should be directed towards relaxing some of the acknowledged limitations of the current modeling. For example, allowing for heterogeneity in the variable estimates across countries would be desirable rather than pooling estimates across countries, as would the use of sub-national data. The analysis of vaccinations should extended once data is available for higher levels of vaccination to properly estimate the likely nonlinear effect beyond the currently low levels reported in this dataset.

Finally, the conclusions reached herein must be placed within the context of and validated by other methodological approaches, such as SIR and agent-based models. However, I have presented significant evidence that strict NPIs provide no further benefits over less stringent ones, and that the latter function primarily as signals for significant voluntary changes in behavior rather than mandatory changes.

## Data Availability

The data is freely available from the following online repositories:
Covid-19 Data Hub Guidotti E and Ardia D. COVID-19 Data Hub. Journal of Open Source Software 2020; 5 (51)2376 34.
Oxford COVID-19 Government Response Tracker Hale T, Angrist N, Goldszmidt R, Kira B, Petherick A, Phillips T, Webster S, Cameron-Blake E, Hallas L, Majumdar S and Tatlow H. A global panel database of pandemic policies (Oxford COVID-19 Government Response Tracker). Nature Human Behaviour 2021 35.
Google Community Mobility Report Google LLC. Google COVID-19 Community Mobility Reports 2020. Available from: https://www.google.com/covid19/mobility/

## Appendix

## Footnotes

1 Unfortunately, while very interesting, a more rigorous game-theoretic analysis would require significantly more data to effectively infer or observe expectations of agents and the multitude of ways to react to said information.

2 Note, if anything this underestimates the possible range of non-significantly different SI ranges, as it ignores the uncertainty associated with whether the range 61–70 is truly the most effective range.

3 I infer that the restriction on gatherings of 100-1000 people is not the main driver of mandatory behavioral change for the following reason. The median level of restrictions on gatherings for the immediately less stringent range, 21–30, is zero, i.e., no restrictions whatsoever. Yet the mean estimate of NPI impact for the 31–40 range is -12.945 compared to -10.173 for the 21–30 range—some of this difference will also be due to other policies that are stricter on average in the former compared to the latter.

4 That is, comparing the baseline of no testing to the first level *χ*^2^ (1) = 12.42, *p* = 0.0013, the first to the second level (*χ*^2^ (1) = 38.55, *p <* 0001) and the second to the third level (*χ*^2^ (1) = 7.83, *p* = 0.0153).

